# Women’s perspectives on fetal movement monitoring in high and low stillbirth settings: a qualitative study

**DOI:** 10.1101/2025.11.12.25340103

**Authors:** Nola Dubuisson, Monica Diez Campa, Abhishek K. Ghosh, Fionnuala M McAuliffe, Niamh C. Nowlan

**Affiliations:** School of Mechanical and Materials Engineering, University College Dublin, Dublin, Ireland; UCD Conway Institute, University College Dublin, Dublin, Ireland; Department of Robotics and Mechatronics Engineering, University of Dhaka, Dhaka 1000, Bangladesh; UCD Perinatal Research Centre, School of Medicine, University College Dublin, National Maternity Hospital, Dublin 2, Ireland

**Keywords:** Fetal Movement Monitoring, Stillbirth, Pregnancy, Qualitative Study, Survey, Interview

## Abstract

**Introduction:** Maternal fetal movement monitoring during pregnancy is commonly advised to assess fetal wellbeing. However, qualitative research exploring how women perceive and implement such advice is lacking, particularly in regions with the highest burden of stillbirth. This study investigates women’s experiences and opinions on fetal movement monitoring during pregnancy across high- and low-stillbirth settings.

**Methods:** Women from three countries with low-stillbirth rates and five with high rates of stillbirth were surveyed and interviewed regarding their experiences with fetal movement monitoring advice. Open-ended answers from the surveys and interview were analysed using inductive thematic methods, while categorical answers were examined using non-parametric statistical analysis.

**Results:** 234 women were included in the study. The nature and extent of fetal movement monitoring advice varied considerably by country, encompassing active monitoring methods such as kick counting, pattern awareness and movement presence detection, as well as guidance on responding to fetal movements concerns. Notably, 33% (37/112) of women in high-stillbirth countries and 8% (10/122) in low-stillbirth countries reported a lack of any fetal movement monitoring advice. Globally, half of women rated the advice easy to follow, while one-third experienced difficulty understanding their healthcare provider’s guidance. Key facilitators of following advice included having an active baby and a clear understanding and confidence in the received advice, whereas barriers included a lack of clarity and understanding, difficulty perceiving movements, competing time demands and challenges in identifying patterns of movements. Maternal anxiety was prevalent, with 78% of participants reporting at least occasional anxiety about fetal movements during pregnancy.

**Conclusion:** Wide variation in the type and consistency of fetal movement monitoring advice across countries indicates the need for further research into the comparative effectiveness of current recommendations, particularly in high-stillbirth settings. High rates of maternal anxiety worldwide highlight the importance of providing support to women navigating fetal movement monitoring.

## 1 Introduction

Stillbirth remains a persistent global health challenge, with an estimated 1.9 million cases occurring in 2023, equivalent to 14.3 stillbirths per 1000 total births worldwide [1]. It is an issue that disproportionally affects low-resource settings, with sub-Saharan Africa and South Asia accounting for 77·4% stillbirths of the global total [2]. Notably, the worst affected country has 20 times higher risk than the country with the lowest stillbirth rate (SBR) [3].

Self-monitoring of fetal movement (FM) is widely advised as a method to monitor fetal well-being and provide an early warning signal for stillbirth risk. Changes or reductions in movements as sensed by the expectant mother have been associated with fetal growth restriction [4, 5], placental abnormalities [6], oligohydramnios [7] and stillbirth [8]. In a study by Warland et al., 63% of women perceived a change in FM before experiencing a stillbirth [9].

Despite the link between reduced FM and stillbirth occurrence, there is currently no universally accepted or standardised method of FM monitoring. Clinical advice given about how and when to monitor movements is inconsistent both between and within countries [10–15]. Large randomised controlled trials (RCTs) and systematic reviews have evaluated a range of FM monitoring strategies, including fetal kick counting [16, 17], increasing maternal awareness of FM with standardised clinical management [18] and the Mindfetalness method which encourages consistent awareness of FM intensity, frequency and character [19]. However, a recent meta-analysis by Hayes et al. (2023) found that there was no definitive proof that any of the FM monitoring approaches reduced stillbirth. A key limitation of prior large-scale RCTs is that they were conducted in high-income settings with a low SBR, and therefore most studies were underpowered to detect the relatively rare outcome of a stillbirth and lacked generalisability to a high-burden settings [20]. In the absence of conclusive evidence, clinical advice about and approaches to FM monitoring vary between different healthcare providers and systems.

Variation in advice provided on FM monitoring in Australia was highlighted by Raynes et al. (2013). They found that several different themes of advice were provided to women, varying from being told about being aware of the normal pattern of movement, to noticing the presence of movement rather than the quantity, to formal kick counting. A number of qualitative studies have sought to understand the experiences of pregnant women engaging with FM monitoring advice. McArdle et al. (2015) surveyed pregnant women in Sydney, Australia and found that they preferred to be given as much information as possible about fetal movements, and that they favoured receiving advice directly from a healthcare professional [12]. Linde et al. (2016) used modified content analysis to determine what types of perceived FM changes prompted women to attend hospital with concerns. Women reported that feeling decreased, absent, weaker, slower and/or a changed pattern of movement would prompt them to seek healthcare advice [11]. Akselsson et al. studied women’s experiences of using the Mindfetalness method and found that the majority of women were positive about the method, with reports of decreased worry, relaxation, creating a connection and increased knowledge of the unborn baby. Some participants expressed that they didn’t have the time to monitor, or that they had no need for the method since their baby was regularly active [21]. A qualitative study by Smyth et al. described the barriers women faced when seeking healthcare support when concerned about FMs and found that participants were worried about being taken seriously, feared interventions such as induction of labour, or had received inconsistent and variable advice about the definition of ‘normal’ movements [14]. AlAmri and Smith conducted a meta-analysis investigating the effect of kick counting on anxiety and fetal-maternal attachment, and found that kick counting did not increase anxiety and promoted fetal-maternal attachment [22].

Although LMICs account for 98.1% of the world’s stillbirths [23], almost no qualitative studies on FM monitoring have been conducted in these settings [10]. A recent review by Dube et al. (2022) surveyed the reported views and experiences of pregnant women in low-income settings relating to reduced FM. Only four relevant studies were identified, none of which were qualitative. We are aware of only one qualitative study focused on the experiences of pregnant women in LMICs, which was published in 2023 [24]. Weller et al. studied perceptions, knowledge and practices of FM self-monitoring among women and healthcare providers in Zanzibar, Tanzania. It found that women often did not know how to monitor FM or when to report concerns, but had an instinctual awareness of their baby’s movement patterns. FM assessment was not routinely discussed in clinical practice, and no protocol was in place for the management of abnormal FM [24].

The majority of studies focused on FM monitoring are based in a single country or setting, and thus a comprehensive comparative cross-country analysis of women’s experiences of FM monitoring remains to be explored. This study addresses this gap, exploring women’s knowledge and experience of following FM monitoring advice across diverse global settings. Specifically, it investigates the types of FM monitoring pregnant women in eight countries in high- and low-stillbirth settings are advised to perform, the ease of following advice, the barriers and facilitators to adhering to the advice, and their experiences of anxiety while monitoring movements.

## 2 Methods

### 2.1 Participants and recruitment

Participants were mainly recruited through worldwide social media advertisements between December 2024 and April 2025. Participants were also recruited through posters in the National Maternity Hospital in Dublin, Ireland. Women were eligible to participate if they were in the third trimester of pregnancy, or if they had given birth within the last year. In addition, participants were required to be over 18 years of age and have a sufficient level of English proficiency to understand and complete the survey or interview. For study integrity and consistency, only participants who had answered at least 80% of the key domain survey questions were included in the final analysis of the survey data. Survey participants indicated in their responses whether they wished to participate in an optional follow-up interview.

Social media (Meta: Facebook and Instagram) advertisements were used to target a sample of countries that either had a high or low SBR according to WHO data [25]. Stillbirth was defined as a baby who died after 28 weeks of pregnancy before or during birth, in accordance with the WHO study on stillbirth [3]. A country was defined as having a high SBR if there were more than 12 stillbirths per 1000 births, in accordance with the WHO Every Newborn Action Plan (ENAP) goal for 2030, while all the low SBR countries included had fewer than 3 stillbirths per 1000 births. To ensure generalisability and a large enough sample population, online targeting focused on countries that had a high proportion of English speakers and people with internet access. The final selection of countries included in the study were chosen based on recruitment success rates of more than 5 participants per country. The low SBR countries included in the analysis were Ireland, the USA and the United Kingdom. The high SBR countries included were Kenya, South Africa, Zimbabwe, Ghana and Nigeria. Online targeted advertising concluded when the number of participants was roughly equivalent between the low and high SBR regions.

### 2.2 Ethics

Ethical approval for this study was granted by the Human Research Ethics Committee in University College Dublin (Ref: LS-C-24-317-Nowlan and LS-25-03Nowlan). Participants were provided with an information sheet prior to accessing the survey and they were required to provide informed consent by clicking “I consent to taking part in the study” before continuing with the survey. A separate information sheet and consent form were provided to the participants before interviewing. All responses were voluntary and no incentives were offered. Participants could skip questions if they wished and could withdraw at any point.

### 2.3 Survey & Interview development

The survey and interview questions were developed for the study, informed by existing literature and clinical guidance. The survey consisted of both categorical and open-ended questions designed to gain an understanding of pregnant women’s experiences and perspectives of monitoring their baby’s movements. Demographic information (pregnancy status, age, parity, country), presence of pregnancy complications or risk factors, and history of miscarriage or stillbirth were collected. Key domain questions were asked about the advice they received about monitoring their baby’s movements, how easy or hard they found it to follow the advice they were given on monitoring their baby’s movements and why, and how often they felt worried or anxious about their baby’s movements. The full list of survey and interview questions can be found in Supplementary Section 2 and Section 3.

### 2.4 Data analysis

Data analyses were conducted using Microsoft Excel 365 (Version 2504), IBM SPSS Statistics (Version 29) and NVIVO (Release 1.7). Frequencies and proportions were used to describe participant demographics and categorical survey questions. Categorical survey questions were compared across demographic subgroups and the statistical effect of demographic parameters on the variable was assessed using either Mann-Whitney tests (for cases of 2 independent groups) or Kruskal-Wallis tests (for k independent groups).

Qualitative responses to open-ended questions were studied using inductive thematic analysis, based on the principles outlined by Braun and Clarke (2006) [26]. The analytical process started with intensive familiarisation of the data, allowing for the development of an initial coding framework by the primary researcher (ND) based on emerging concepts and recurring patterns. The coding framework was developed through multiple iterations and through discussion with the research team.

A subset of 100 participant responses (42% of the total dataset) was then independently coded by a second researcher (MDC), who was blinded to the initial coding. Discrepancies in the coding were identified and resolved through consensus-based discussion between the two coders and a third member of the research team (NCN). During this iterative process, the coding framework was further refined to clarify code definitions. The remaining responses were then coded using the new framework by ND, ensuring consistency across the entire dataset. Finally, the refined codes were iteratively grouped into broader themes. Response quotes that demonstrated the themes were selected and tagged according to whether they were from the survey (S) or from the interview (I), with the participant number and country.

Quantitative findings were strategically used to provide context and identify patterns in the qualitative responses. Frequencies of qualitative themes were compared between settings to understand notable patterns and to highlight prominence, but do not imply statistical significance.

## 3 Results

A total of 359 women responded to the survey, of which 234 were included in the study. Of those excluded, 111 did not answer the required number of questions, two provided incomplete or nonsensical answers, seven did not specify a country, four were from countries with fewer than five participants, and one was not yet in the third trimester. Fifteen of the included participants opted in for a follow-up interview.

The demographics of the survey and interview participants are shown in Table 1. Survey participants were recruited to be evenly distributed between high (*n* = 112) and low (*n* = 122) stillbirth rate countries. Most survey participants were currently pregnant (66.7%), between 30-39 years of age (64.1%) and multiparous (56.8%). Interviews had more participants from countries with a low SBR (80%).

**Table 1:** Maternal characteristics.

| Characteristic | Survey<br>(n=234) | Interview<br>(n=15) |
| --- | --- | --- |
| Pregnancy status |  |  |
| Currently | 156 (66.7%) | 5 (33.3%) |
| Recently | 78 (33.3%) | 10 (66.7%) |
| Country stillbirth rate |  |  |
| High | 112 (47.9%) | 3 (20.0%) |
| Low | 122 (52.1%) | 12 (80.0%) |
| Country |  |  |
| Kenya (High SBR) | 18 (7.7%) | 1 (6.7%) |
| South Africa (High SBR) | 55 (23.5%) | – |
| Zimbabwe (High SBR) | 26 (11.1%) | 1 (6.7%) |
| Other* (High SBR) | 13 (5.6%) | 1 (6.7%) |
| Ireland (Low SBR) | 47 (20.1%) | 4 (26.7%) |
| UK (Low SBR) | 36 (15.4%) | 1 (6.7%) |
| USA (Low SBR) | 39 (16.7%) | 7 (46.7%) |
| Age group (years) |  |  |
| 18–29 | 64 (27.4%) | – |
| 30–39 | 150 (64.1%) | – |
| 40–49 | 20 (8.5%) | – |
| Parity |  |  |
| Primiparous | 101 (43.2%) | 5 (26.7%) |
| Multiparous | 133 (56.8%) | 9 (60.0%) |
| Undisclosed | – | 1 (6.7%) |
| Complication/risk factor |  |  |
| Yes | 146 (62.4%) | – |
| No | 88 (37.6%) | – |
| Prior miscarriage |  |  |
| Yes | 63 (26.9%) | – |
| No | 171 (73.1%) | – |
| Prior stillbirth |  |  |
| Yes | 14 (6.0%) | – |
| No | 218 (93.2%) | – |
| Undisclosed | 2 (0.9%) | – |

\*"Other" countries in the survey (with <10 respondents) included Ghana (n=7) and Nigeria (n=6). One interview participant was from Nigeria.

Pregnancy complications and/or risk factors were reported by more than half of survey participants (62.6%). Approximately a quarter of survey participants reported having a prior miscarriage (27.2%) and 14 participants (6.0%) reported having a prior stillbirth. Interview participants were not asked about their age, pregnancy complications or prior losses.

### 3.1 Movement monitoring advice

We identified three categories of the type of advice received on FM monitoring, namely a) active monitoring; b) reactive monitoring (when and how to act if concerned about FM) and c) no advice.

#### Active monitoring advice

Participants reported a variety of advice about how to keep track of FMs, including counting kicks, tracking patterns of movement, and checking periodically for the presence of movements.

Participants described being advised to do kick counting to assess the frequency of FMs, often with the aim of reaching a defined number of kicks within a certain time period. The expected number of kicks and the prescribed timing varied between responses. A count of 10 kicks in 2 hours was the most commonly advised guideline. However, different participants mentioned counts of 10 kicks per day, 20 kicks per day, 10 kicks per hour and 20 kicks per hour. As shown in Figure 1, kick counting was mentioned by a few participants in every country, but it was most prevalent in the USA with 59% of participants referencing it in their responses. Although described by fewer participants overall, kick counting was also one of the more common types of advice in South Africa.

**Figure 1:**
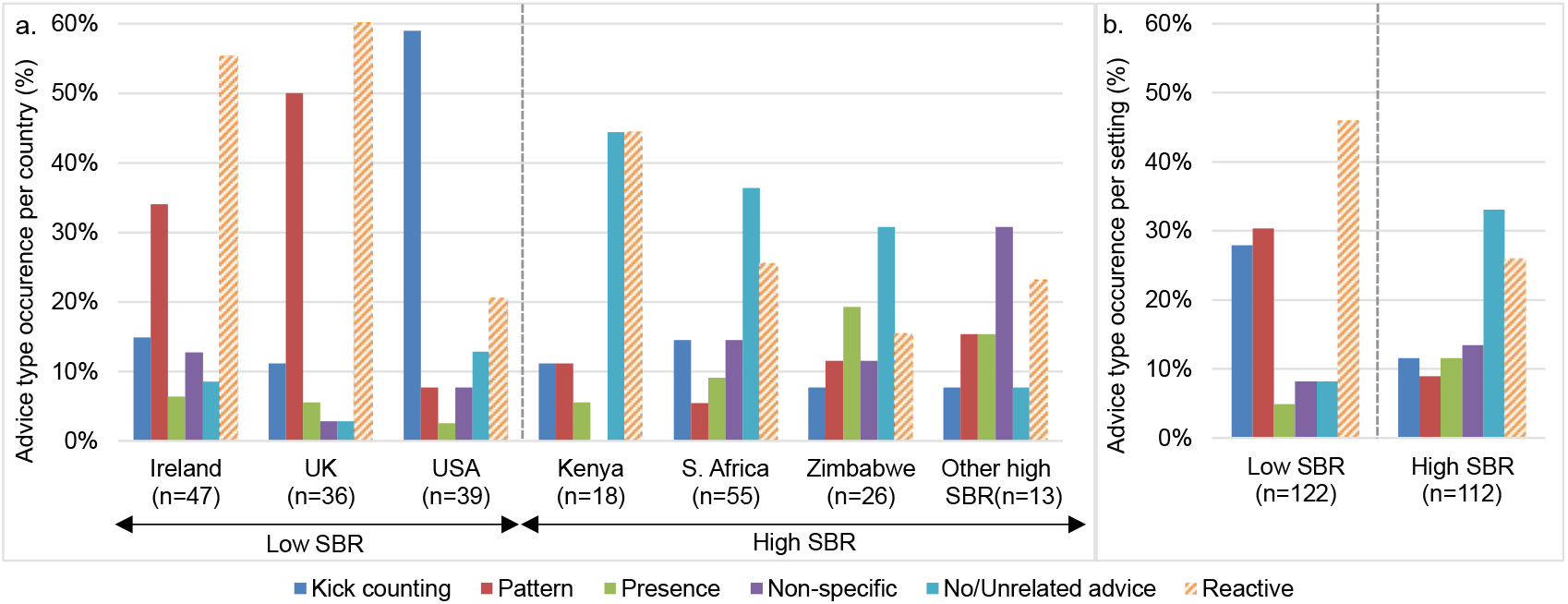
FM monitoring advice varied between countries and settings. 31-44% of participants from high SBR countries had received no/unrelated advice, compared to *<*15% of participants from low SBR countries. Pattern based monitoring was predominantly advised in Ireland and the UK, while kick counting was more predominant in the USA. Kick counting, pattern recognition and presence-based monitoring were all advised to a similar extent in high SBR countries, with higher rates of kick counting advice in South Africa and of presence-based monitoring in Zimbabwe. Reactive advice about when and how to act was particularly emphasised in Ireland, the UK and Kenya.

> “*Every day, pick the same time, and in the 2 hours, count movement at least 10 times*.*” (S69, USA)*

Pattern tracking was described by participants as learning the baby’s normal movement routine and being aware of changes. It was the most commonly described method in Ireland and the UK, with approximately a third of Irish and half of UK participants mentioning pattern tracking. Pattern tracking was also reported by some participants in every high SBR country as a method of FM monitoring.

> “*Monitor over the next 6 weeks for a pattern and track to make sure baby stays in that pattern*” *(S195, UK)*

Detecting the presence of movement regularly was described by several participants, with time frames ranging from hourly to daily. Unlike kick counting, this advice did not involve counting movements but focused only on confirming that movement had occurred. Presence-based advice was a more predominant method in high SBR countries than in low SBR countries. It was the most common form of advice in Zimbabwe, described by approximately a fifth of participants.

> “*Listen 3 to 4 times a day if the baby is moving*” *(S114, Zimbabwe)*

Several participants described receiving advice about monitoring movements, but their responses did not specify what type of monitoring they were instructed to do. These cases are labelled as non-specific in Figure 1.

In low SBR countries, advice type was dominated by one category of monitoring method, with pattern recognition dominating in Ireland and the UK, and kick counting dominating in the US. In contrast, there was less coherence in the advice received in high SBR countries, with advice types split between kick counting, pattern recognition, presence detection, as shown in Figure 1.

#### Reactive monitoring advice (when and how to act)

Advice about what to watch out for as warning signs of fetal distress was often described in participant responses, often reported alongside other forms of advice. A reduction, absence or change in movements were the types of warning signs most commonly advised. An increase in movements was only referenced by two participants, one each from Ireland and the UK. Maternal concern or intuition was sometimes referenced by participants as a trigger for seeking care, especially in Ireland and the UK.

> “*Trust your gut. If you are worried at all attend (*…*) hospital*” *(S21, Ireland)*

Reactive monitoring advice was particularly emphasised in Ireland and the UK, reported in over half of responses. UK and Irish participants most commonly referred to reduced or changed movements as a reason to seek care, but absence of movements and general maternal concern were also mentioned.

> “*Get in touch with the Midwife team if I notice any changes in Baby’s movements. This includes them being quieter/less active than normal, but also if they’re being more active than normal*.” *(S89, UK)*

Most participants from the USA did not report being told about warning signs. Of the small number who did, changes in movements were most frequently referenced. A large proportion of Kenyan participants were advised about warning signs, in particular with a focus on the reduction or absence of movements. In South Africa and Zimbabwe, the warning signs were almost exclusively reported as a complete absence of movement, with time periods ranging from no movements in several hours to none in a day.

> “*If your baby doesn’t move in 24 hours, visit the hospital*” *(S170, South Africa)*

As illustrated in the quote above, participants often described being instructed on what actions to take if they were concerned about movements. Over a quarter of all participants (27%) explicitly mentioned being told to seek medical care, by calling their midwife or doctor, or by going into the hospital directly. This was particularly emphasised in Ireland, the UK and Kenya, while only a few participants from the other high SBR countries stated explicitly that they were told to seek medical care. In contrast to other low SBR countries, only three participants from the USA (8%) explicitly describe being advised to seek medical care for concerns about FMs.

A few participants described being told to postpone seeking direct care by trying to stimulate movement first and monitoring for resulting movements. Stimulation strategies included drinking or eating something and lying on one side. Overall this type of advice was not common, with the highest occurrence of it in Ireland where 9% of participants referred to it.

> “*If no movements felt, drink a fizzy drink/eat something & wait half an hour to see if response*” *(S25, Ireland)*

#### No advice

Some participants described getting no advice on FM monitoring in their responses. In other instances, when specifically asked what type of advice they were given about FM monitoring, some participants would describe advice that was unrelated to movements, possibly indicating they were not aware of the concept.

> “*Very little to none*” *(S36, Ireland)*
>
> “*How to sit and sleep* …*exercises to do, what to eat and what not to eat*” *(S223, South Africa)*

Between the two types of responses, a fifth of participants responses indicated a lack of sufficient advice. This was particularly prevalent in countries with a high stillbirth rate, with between 31-44% of participants from Kenya, South Africa and Zimbabwe indicating a lack of advice compared to 3-13% in low SBR countries, as shown in Figure 1. The UK had the lowest incidence of receiving no/unrelated advice, while South Africa had the highest.

#### 3.1.1 Ease of following advice

Approximately half of the participants (115/234) found it easy to follow advice (very easy/easy/somewhat easy), while about a third (74/234) reported some degree of difficulty (very difficult/difficult/somewhat difficult).

Participants from countries with a low SBR found it significantly more difficult to follow advice compared to participants from countries with a high SBR (*p <* 0.001), as shown in Supplementary Table 2. There was also a significant difference in the rating of ease of following advice between individual countries (*p* = 0.017), although no significant pairwise groupings were found between countries. As seen in Figure 2, Irish, UK and South African participants found it most difficult to follow advice, while participants from the USA, Kenya, and ‘Other high SBR’ countries found it easiest.

**Figure 2:**
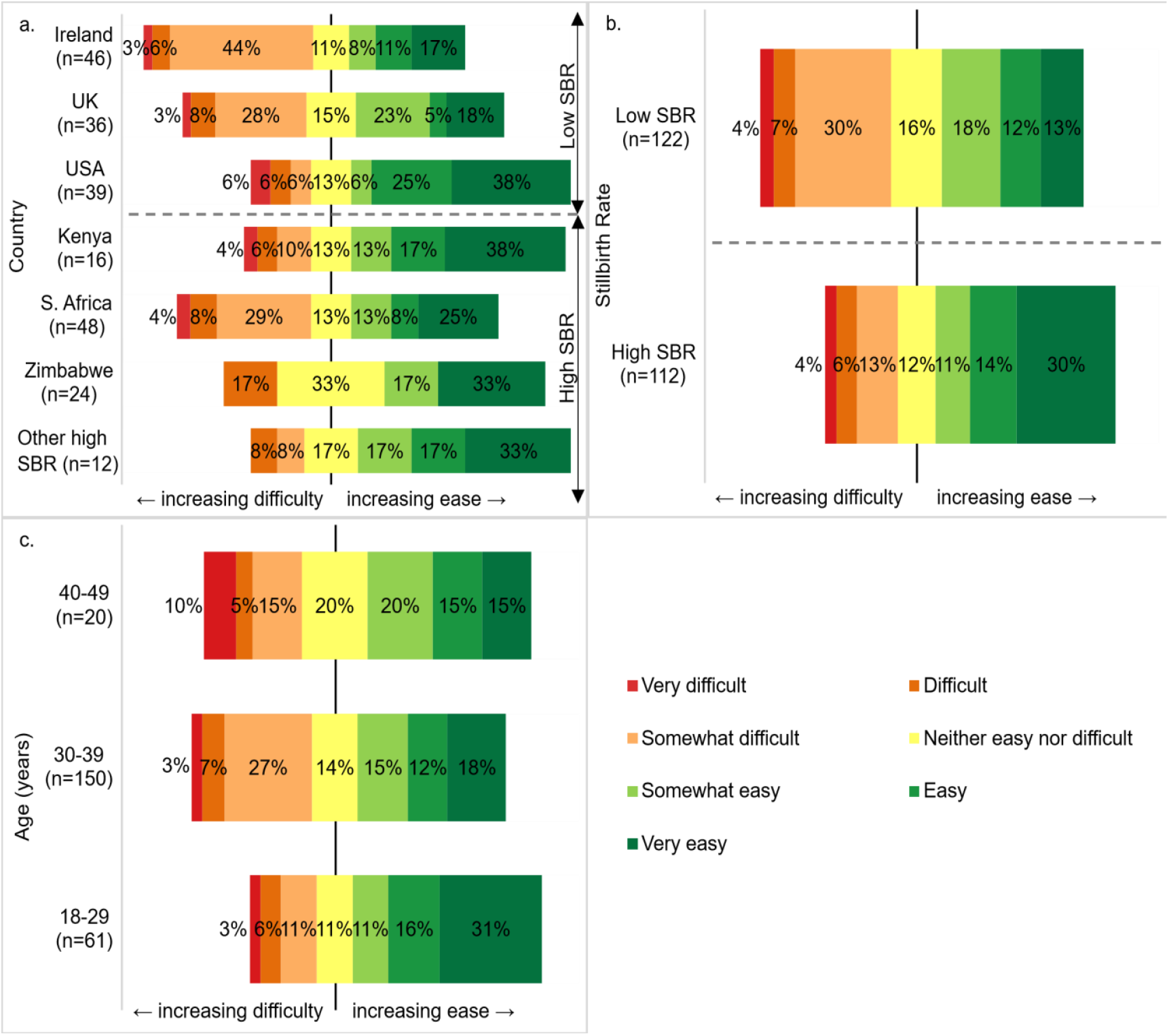
(a) Reported ease of following FM monitoring advice was significantly higher in high stillbirth countries than in low stillbirth countries. (b) There was also a significant difference in ease between countries, however no pairwise groupings were significant. (c) Participants aged 18-29 years found it significantly easier to follow advice than participants aged 30-39 years. No other factors had a significance effect on reported ease of following advice.

Age was also shown to have a significant impact on ease of following advice (*p* = 0.030), with participants aged 18-29 years reporting significantly greater ease than those aged 30-39 years, as seen in Figure 2. While not statistically significant, trends were observed suggesting that primiparous participants, those with pregnancy complications or risk factors, and those with a history of prior miscarriage or stillbirth tended to report greater difficulty with following advice.

#### Facilitators

Two main factors were identified in participant responses as reasons for increased ease of following advice: firstly, having a clear understanding and confidence in monitoring, and secondly, having an active baby.

Having a clear understanding of how to monitor FMs and what to expect was identified as a factor that made monitoring easier by 19% of survey participants. This was facilitated by clear instructions from medical professionals, knowledge from a prior pregnancy, and/or a strong confidence in their own understanding of what to expect and how to identify reduced movements, as described by the following participants. Participants from high SBR countries described having a clear understanding and confidence in the advice more frequently than low SBR countries.

> “*Information was included in hospital notes and highlighted by midwife during appointment. Movement was discussed at each appointment to reiterate information*” *(S135, UK)*
>
> “*Well since I was pregnant before I go on what I know*” *(S135, South Africa)*

Having an active baby who moved often made monitoring easier for women in both high and low SBR settings, particularly in the USA, South Africa, Zimbabwe and ‘Other Countries’. An active baby made it easier to keep track of movements, gave reassurance that the baby was healthy, and increased confidence in being able to identify changes easily.

> “*My baby was a very active baby so I definitely would have been able to pick up any slightest change*.” *(S41, UK)*

#### Barriers

The primary themes identified by participants who reported difficulty in following FM monitoring advice were 1) lack of clarity and understanding; 2) struggling to identify a pattern; 3) difficulty perceiving movements; and 4) competing time demands.

The most frequently observed challenge participants reported was a lack of clarity and understanding about self-monitoring of FM. This was especially prevalent in low SBR countries with 28% of participants indicating it in their responses compared to 12% of participants from high SBR countries. Participants expressed that they felt the advice was insufficient, that they didn’t know what to expect, and were uncertain about when to worry about their baby’s movements. In Ireland and the UK, where an emphasis is placed on identifying “normal” movement rhythms, as described in Section 3.1, participants expressed that they did not know what normal or healthy movement should be. This was described by an interview participant from the UK:

> “*It’s very difficult to know what’s normal. And when you ask a professional, they basically say: “we can’t tell you, because everybody’s different”, which I appreciate, but it does leave a lot of room for interpretation, so it is very unsettling*.” *(I11, UK)*

Struggling to identify a pattern in their baby’s movement was frequently reported by Irish and UK participants, but also by some participants in the USA and Zimbabwe. Participants expressed that they found it hard to recognise a pattern in their baby’s movements, or that variations in their baby’s movements made monitoring more difficult, as described by this Irish participant:

> “*I don’t feel my baby has developed a pattern and is quieter some days and more active others*” *(S40, Ireland)*

Another challenge experienced by participants was difficulty perceiving movements or having a “quiet baby”. Participants sometimes attributed this to their anterior placenta, which can cushion the fetus and reduce the ability to feel movements clearly. Others found it difficult when their babies were quiet or had periods of reduced movement, which often contributed to anxiety about their baby’s wellbeing. As an interviewee from Zimbabwe said:

> *“It’s [the advice] not easy to follow, because you can’t really tell if the fetus is moving, or if it’s not sometimes, you can’t tell if your baby is alive or not, or if your baby is in distress, you can’t tell*.*” (I13, Zimbabwe)*

Half of the interview participants and 14% of survey participants expressed that other demands on their time, such as work, childcare or other responsibilities, made it more difficult to keep track of movements. This was observed as a challenge that was universal across most countries; however, it was particularly prevalent in the USA, with 23% of survey participants identifying it as an issue. For example, a participant said:

> *“I also have another child and it’s hard to sit and relax to count them, I’m often busy and don’t remember the last time I felt her move*.*” (S76, USA)*

As reported in Section 3.1, many survey participants reported receiving no or unrelated information about FM monitoring. In such cases, most participants answered “n/a” or “neither easy or difficult” to the question about ease of following FM monitoring advice. Although the absence of advice generally did not contribute to the difficulty rating, it represents a clear barrier to the overall effectiveness of FM monitoring.

### 3.2 Anxiety about fetal movements

The majority of participants (78%) reported feeling anxious or worried about their baby’s movements at least some of the time (24% always, 14% usually and 40% sometimes), while the remainder felt anxious rarely (16%) or never (6%).

Participants from countries with a high stillbirth rate were significantly more likely to report anxiety about their baby’s movements than participants from countries with a low stillbirth rate (p=0.039). Although the difference between individual countries was not significant, South Africa and Kenya had the highest proportion of participants reporting always or usually anxious, while Kenya and the USA had the highest levels of participants who were “Always” anxious, as seen in Figure 4. Factors other than location that significantly impacted frequency of anxiety were pregnancy complications or risk factors (p=0.005), and experience of a prior stillbirth (p=0.012), as shown in Supplementary Table 4. The effects of pregnancy complications or the experience of a prior loss were also noted as a source of anxiety by several of the interview participants.

**Figure 3:**
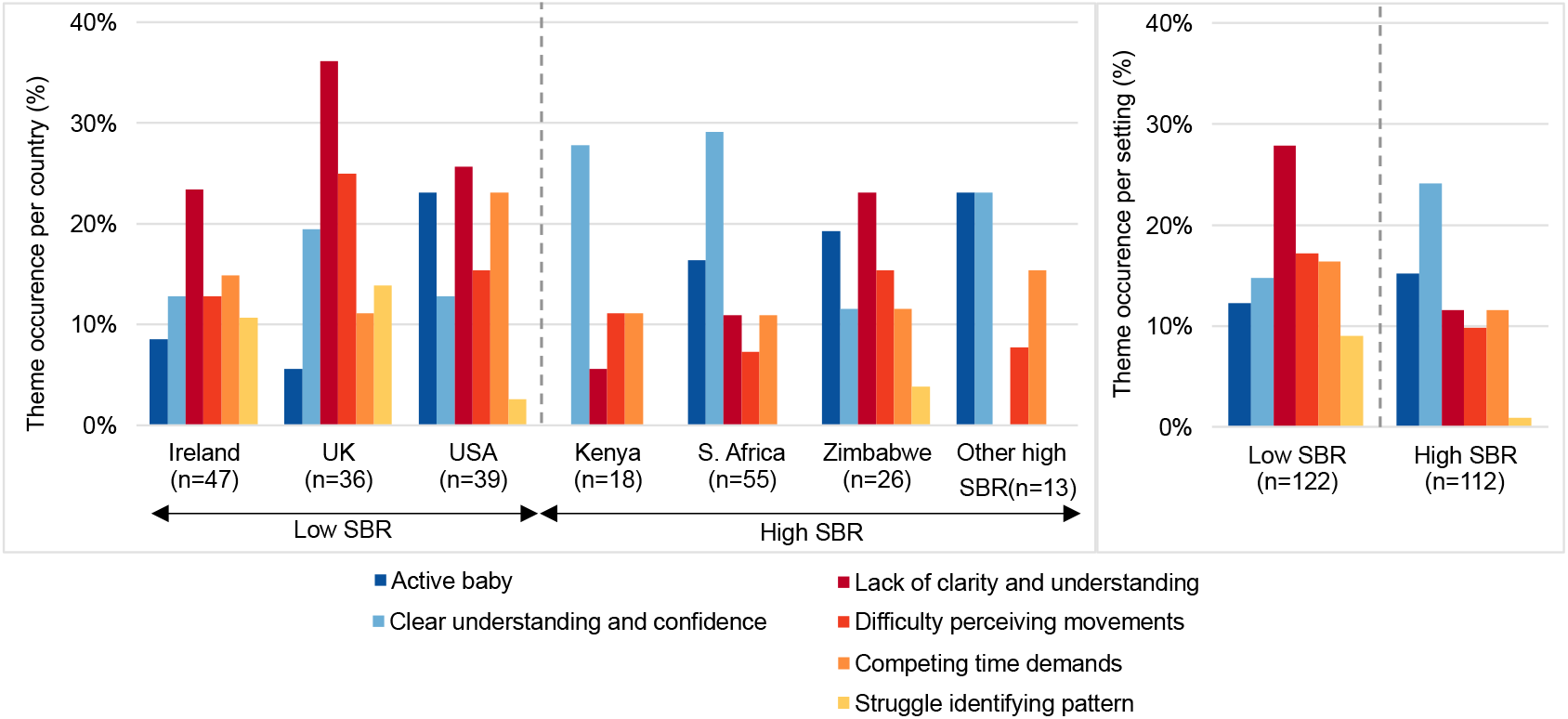
Across most countries, participants who could easily feel their baby moving found it easier to follow advice, while participants who had difficulty perceiving movements found it harder. Having a clear understanding of and confidence in received advice contributed to higher ease in monitoring in high SBR countries, while lack of clarity and understanding contributed to higher difficulty in low SBR countries. A common barrier across all countries was competing time demands, while struggling to identify a consistent pattern was a barrier in countries which advised pattern-based monitoring.

**Figure 4:**
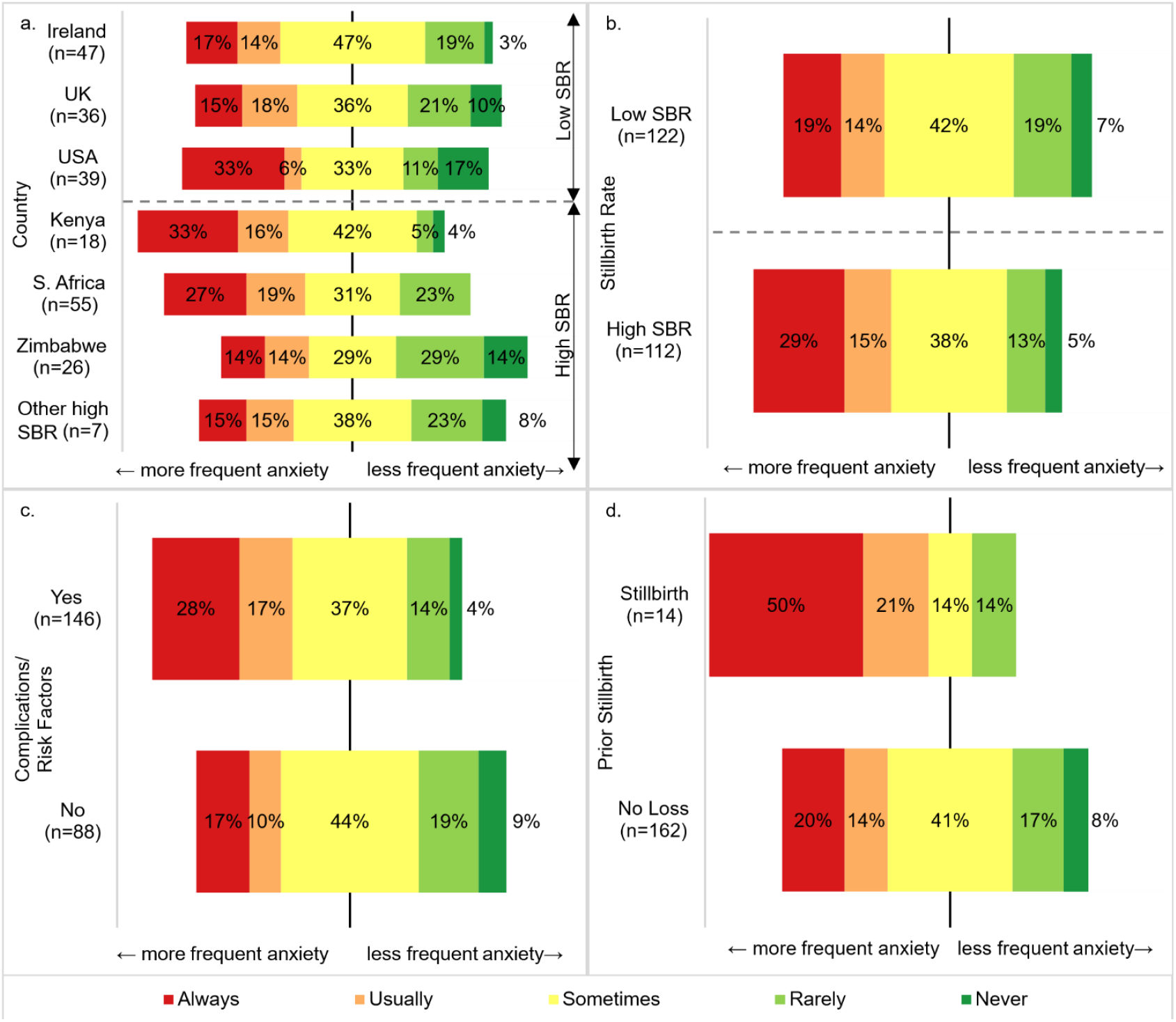
(a) There was no significant difference between ratings of anxiety frequency between countries, however some trend towards more or less frequent anxiety. (b) Participants from high SBR countries are significantly more frequently anxious about movements than those from low SBR countries. (c) Participants with pregnancy complications/risk factors generally rated their frequency of anxiety as higher than those without. (d) Participants with experience of a prior stillbirth were significantly more frequently anxious than those who hadn’t.

> “*Yes, with my first I did [feel anxious] a lot because I had complications, and one of them was [obstetric] cholestasis, which has the increased risk of stillbirth, so I was always convinced that if I didn’t feel her for a long time, that she was like actively passing away*.” *(I6, USA)*
>
> “*I was actually very anxious, especially after losing the 1st pregnancy*.” *(I10, Kenya)*

Parity, age and experience of a prior miscarriage were not shown to have a significant effect on anxiety, although women between the ages of 18-29 and those with a prior miscarriage trended towards more frequent anxiety than other participants.

### 3.3 Synthesis across countries and settings

The countries where participants found it hardest to follow advice on fetal activity monitoring were Ireland, the UK and South Africa. The country-specific themes most associated with difficulty in following advice were lack of clarity and understanding (Ireland, UK) and difficulty in perceiving movements (UK). No predominant theme emerged from the reports of the South African participants to provide insight into the challenges experienced by women there in following the advice. The USA, Kenya and ‘Other high SBR’ countries found it easiest to follow FM monitoring advice. The themes that were most associated with countries that found following advice easiest were having an active baby (USA) and having a clear understanding and confidence in the advice (Kenya). Our findings suggest that some types of advice may be easier to follow than others. Given the ratings of difficulty in Ireland and the UK, pattern recognition monitoring may be more difficult for women to follow, while the results from the USA imply that kick counting might be easier to follow.

Participants in high-SBR countries reported receiving no advice or unrelated advice more commonly than in low-SBR countries. Furthermore, fewer participants in high-SBR countries reported receiving active monitoring advice (kick counting/pattern recognition/presence) than in low-SBR countries. An interesting difference between the included high- and low-SBR countries is that in each of the low-SBR countries there was a dominant theme of advice (either pattern recognition or kick counting, Figure 1), while none of the high-SBR countries exhibited a dominant type of “active” monitoring advice (Figure 1).

Women in high-SBR countries were more likely to report a clear understanding and confidence in FM monitoring than those in low-SBR countries, with the exception of Zimbabwe. Zimbabwe participants reported “lack of clarity and understanding” more than any other high-SBR country, but also reported the lowest rates of anxiety among the high-SBR countries. Zimbabwe did not stand out from other high-SBR countries in terms of the types of advice received, except for a slight decrease in the proportion of “reactive advice” given in Zimbabwe compared to other high-SBR countries.

Kenya and the USA had the highest incidences of “always” anxious women, while simultaneously being the countries that reported the greatest ease of following advice. Both countries have vastly different SBRs [25], and different types of advice received (Figure 1). This finding implies that providing advice that is easy to follow does not necessarily provide reassurance in terms of anxiety about FMs.

## 4 Discussion

This study is unique in its inclusion of women from eight different countries, spanning both low and high stillbirth rate settings, allowing comparisons of how advice on FM monitoring is given, interpreted and experienced across diverse settings. In this regard, we observed marked differences between both individual countries and between groups of countries categorised by stillbirth rate. In particular, we found a systemic difference in the coherence of advice messaging between settings. Low SBR countries demonstrated greater consistency in the themes of advice, compared to high SBR countries. Despite such differences, a shared global concern among pregnant women was maternal anxiety about FMs - 78% of women reported feeling anxious about their baby’s movements at least occasionally during their pregnancy. Our findings highlight the importance of clear and consistent advice on FM monitoring and the need for supportive care that acknowledges and eases maternal anxiety.

Coherence of advice was observed as a distinct difference between high and low SBR settings. The type of FM monitoring advice also varied considerably across countries. In low SBR countries, a predominant theme of advice, e.g. kick counting in the USA and FM pattern recognition in Ireland and the UK, were evident. In contrast, women in high SBR countries frequently reported receiving little or no advice, and among the women who did receive advice, the content varied widely. This aligns with previous findings reporting limited awareness of absent or reduced FMs as a warning sign in LMIC, with awareness rates as low as around 4.6% in Ghana [27], 11.7% in Jordan [28] and 16.4% in Nigeria [29]. Differences in the number of midwives and obstetricians across countries may affect the consistency and quantity of advice given to pregnant women. Without standardisation of advice about FM monitoring, particularly given that women may find information on websites from other countries, can lead to a confusing mix of advice from healthcare providers, family and friends and the internet. Such inconsistency risks misinformation, or delayed care as reported by Smyth et al. [14]. One of the barriers to effective FM monitoring identified in our study was a lack of understanding and clarity about advice, to which variation in received advice can contribute. While the standardisation of national guidelines about FM monitoring might be expected to improve clarity for both healthcare providers and pregnant women, our findings suggest otherwise. Participants in the UK [30], Ireland [31], and South Africa [32], countries with national FM monitoring guidelines, were among those who reported the greatest difficulty in following advice. This suggests that the challenge may lie less in the consistency of the advice, and more in the content and delivery of the advice itself.

The variability in the advice received, particularly in high-SBR countries, may be attributable to the absence of conclusive evidence on the effectiveness of FM monitoring. Several large-scale randomised trials have investigated the potential of FM monitoring to reduce stillbirth occurrence, including the AFFIRM trial in the UK [18], the Mindfetalness trial in Sweden [19], and the My Baby’s Movements trial in Australia and New Zealand [33]. However, none of the trials found significant reductions in stillbirth rates. As highlighted by Housseine et al., the global applicability of these trials is limited as these trials were conducted in places with already low stillbirth rates and relatively high maternal awareness of FM monitoring. Consequently, these studies may have been underpowered to detect the relatively rare outcome of stillbirth [34] and may not accurately present the potential benefits of FM monitoring in lower-resource settings. It is imperative that further research is conducted in LMICs to determine whether FM monitoring awareness could be an effective method of reducing stillbirth rates in populations bearing the highest burden of stillbirth.

For FM monitoring methods to be effective, it is important to understand pregnant women’s perspectives, including the facilitators and barriers they experience while trying to follow the advice they are provided. Our study found that participants from countries with a high SBR generally found it easier to follow FM monitoring advice, with the exception of Zimbabwe, participants described that they felt confident in their knowledge of what to expect and that they had a clear understanding of how to monitor. Conversely, participants from low SBR countries and Zimbabwe expressed a higher level of uncertainty and lack of understanding about received advice, in particular when it came to the specifics of how to monitor, when to worry and what was “normal”. These findings align with those of McArdle et al., who reported that pregnant women would prefer to receive as much information as possible [12]. These results suggest that the provision of a clear comprehensive guideline about how to monitor FMs would be beneficial.

Across all settings, a key factor that affected ease of following FM monitoring advice was how frequently participants could feel their fetus moving. Participants who described their fetus as more active found it easier to follow the advice they were given, whereas those who experienced difficulty in perceiving FMs found it harder to follow the FM monitoring advice given. Additionally, participants across all settings reported that being busy with other time demands, such as work or children, made it more difficult to both sense and monitor movements. Strategies to monitor FMs should therefore be considerate of these commonly experienced barriers. Research is ongoing to develop wearable FM monitors, for example [35, 36], but none are yet available commercially.

Across the diverse range of settings in this study, a shared global factor was the high prevalence of maternal anxiety about FMs, with four out of five women reporting experiencing anxiety about FMs at least occasionally. Notably, countries with a higher ease of following advice were not necessarily associated with lower rates of anxiety, indicating that even when advice is easy to follow, anxiety persists. Participants from countries with a high stillbirth rate, and participants with pregnancy complications, risk factors, and experience of a prior stillbirth were significantly more frequently anxious about FMs. These findings are consistent with previous studies that demonstrated elevated levels of anxiety in medically complicated pregnancies [37] and in pregnancies following a stillbirth [38]. Additionally, participants from high SBR settings are more likely to know other people who have had a stillbirth and therefore may have a greater understanding of the importance of FM monitoring. Several studies have explored the effect of kick counting on anxiety, and a meta-analysis by AlAmri and Smith found no difference in maternal anxiety between women who formally counted FMs and those who did not [22]. To our knowledge, there has not been a large-scale study on the effect of pattern-based monitoring or presence-based monitoring on maternal anxiety, or a study comparing the impact of different methods of advice types on anxiety. Since maternal anxiety can impact the health of both the expectant mother and fetus [39, 40], it is important that the impact of FM monitoring guidelines on anxiety should be appropriately investigated and assessed alongside other outcome metrics. Particular care should be taken when providing advice to cohorts who experience higher rates of anxiety about FM monitoring.

There were some limitations to our study. Participation was limited to women who spoke English and had internet access. Additionally, as participants opted in to answer the online survey and interviews, findings may be influenced by self-selection bias. Finally, women in our study were interviewed and surveyed up to a year after giving birth, which may introduce recall bias.

## 5 Conclusion

This study revealed substantial variation in the consistency and type of FM monitoring advice received by women across both high and low stillbirth rate countries, with a high rate of women receiving no advice at all. Our findings highlight the need for clarity and consistency in FM monitoring communication. Further studies on the comparative effectiveness of FM monitoring type are warranted. The research gap regarding FM monitoring is particularly pronounced in places with the highest burden of stillbirth. The high rates of anxiety related to FM reported by women from across the globe indicate a need for further support and sensitivity when designing FM monitoring methods and guidelines.

## Data Availability

All survey data produced in the present study are available on Zenodo.
Interview excerpts are available upon reasonable request to the authors.

https://doi.org/10.5281/zenodo.17572671

## Acknowledgments

This research was funded by the Wellcome Leap In Utero Programme.

## Availability of data and materials

The anonymous survey data is stored on a public repository and can be found at: https://doi.org/10.5281/zenodo.17572671 [41]. Interview transcripts available upon reasonable request.

## References

[1] UNICEF. Stillbirths and Stillbirth Rates;. Available from: https://data.unicef.org/topic/child-survival/stillbirths/.

[2] Comfort H, McHugh TA, Schumacher AE, Harris A, May EA, Paulson KR, et al. Global, Regional, and National Stillbirths at 20 Weeks’ Gestation or Longer in 204 Countries and Territories, 1990–2021: Findings from the Global Burden of Disease Study 2021;404(10466):1955–88.

[3] Arora A. Never Forgotten: The Situation of Stillbirth around the Globe;. Available from: https://data.unicef.org/resources/never-forgotten-stillbirth-estimates-report/.

[4] Tsikouras P, Antsaklis P, Nikolettos K, Kotanidou S, Kritsotaki N, Bothou A, et al. Diagnosis, Prevention, and Management of Fetal Growth Restriction (FGR);14(7):698.

[5] Lu Y, Palin V, Heazell A. Risk Factors for Adverse Pregnancy Outcomes in Reduced Fetal Movement: An IPD Meta-Analysis;132(7):1000–9.

[6] Kuile M, Erwich JJHM, Heazell AEP. Stillbirths Preceded by Reduced Fetal Movements Are More Frequently Associated with Placental Insufficiency: A Retrospective Cohort Study;50(6):668–77.

[7] Padore IS, Jinturkar AA. Study of Obstetric and Fetal Outcomes Associated with Maternal Perception of Decreased Fetal Movements in Third Trimester;13(5):1271–6.

[8] Carroll L, Gallagher L, Smith V. Pregnancy, Birth and Neonatal Outcomes Associated with Reduced Fetal Movements: A Systematic Review and Meta-Analysis of Non-Randomised Studies; 116:103524.

[9] Warland J, O’Brien LM, Heazell AEP, Mitchell EA, the STARS consortium. An International Internet Survey of the Experiences of 1,714 Mothers with a Late Stillbirth: The STARS Cohort Study;15(1):172.

[10] Smith V, Muldoon K, Brady V, Delaney H. Assessing Fetal Movements in Pregnancy: A Qualitative Evidence Synthesis of Women’s Views, Perspectives and Experiences; 21(1):197.

[11] Linde A, Georgsson S, Pettersson K, Holmstro m S, Norberg E, Ra destad I. Fetal Movement in Late Pregnancy – a Content Analysis of Women’s Experiences of How Their Unborn Baby Moved Less or Differently;16(1):127.

[12] McArdle A, Flenady V, Toohill J, Gamble J, Creedy D. How Pregnant Women Learn about Foetal Movements: Sources and Preferences for Information;28(1):54–9.

[13] Raynes-Greenow CH, Gordon A, Li Q, Hyett JA. A Cross-Sectional Study of Maternal Perception of Fetal Movements and Antenatal Advice in a General Pregnant Population, Using a Qualitative Framework;13(1):32.

[14] Smyth RMD, Taylor W, Heazell AE, Furber C, Whitworth M, Lavender T. Women’s and Clinicians Perspectives of Presentation with Reduced Fetal Movements: A Qualitative Study;16(1):280.

[15] Warland J, Glover P. Fetal Movements: What Are We Telling Women?;30(1):23–8.

[16] Bellussi F, Po’ G, Livi A, Saccone G, De Vivo V, Oliver EA, et al. Fetal Movement Counting and Perinatal Mortality: A Systematic Review and Meta-analysis;135(2):453.

[17] Delaram M, Jafarzadeh L. The Effects of Fetal Movement Counting on Pregnancy Outcomes;10(2):SC22–4.

[18] Norman JE, Heazell AEP, Rodriguez A, Weir CJ, Stock SJE, Calderwood CJ, et al. Awareness of Fetal Movements and Care Package to Reduce Fetal Mortality (AFFIRM): A Stepped Wedge, Cluster Randomised Trial;392(10158):1629–38.

[19] Akselsson A, Lindgren H, Georgsson S, Pettersson K, Steineck G, Skokic V, et al. Mindfetalness to Increase Women’s Awareness of Fetal Movements and Pregnancy Outcomes: A Cluster-Randomised Controlled Trial Including 39 865 Women;127(7):829–37.

[20] Hayes DJL, Dumville JC, Walsh T, Higgins LE, Fisher M, Akselsson A, et al. Effect of Encouraging Awareness of Reduced Fetal Movement and Subsequent Clinical Management on Pregnancy Outcome: A Systematic Review and Meta-Analysis; 5(3).

[21] Akselsson A, Georgsson S, Lindgren H, Pettersson K, Ra destad I. Women’s Attitudes, Experiences and Compliance Concerning the Use of Mindfetalness-a Method for Systematic Observation of Fetal Movements in Late Pregnancy;17(1):359.

[22] AlAmri N, Smith V. The Effect of Formal Fetal Movement Counting on Maternal Psychological Outcomes: A Systematic Review and Meta-Analysis; 6:10.

[23] Hug L, You D, Blencowe H, Mishra A, Wang Z, Fix MJ, et al. Global, Regional, and National Estimates and Trends in Stillbirths from 2000 to 2019: A Systematic Assessment;398(10302):772–85.

[24] Weller K, Housseine N, Khamis RS, Meguid T, Hofmeyr GJ, Browne JL, et al. Maternal Perception of Fetal Movements: Views, Knowledge and Practices of Women and Health Providers in a Low-Resource Setting;3(3):e0000887.

[25] WHO. Stillbirth Rate (per 1000 Total Births);. Available from: https://www.who.int/data/gho/data/indicators/indicator-details/GHO/stillbirth-rate-(per-1000-total-births).

[26] Braun V, Clarke V. Using Thematic Analysis in Psychology;3(2):77–101.

[27] Udofia EA, Obed SA, Calys-Tagoe BNL, Nimo KP. Birth and Emergency Planning: A Cross Sectional Survey of Postnatal Women at Korle Bu Teaching Hospital, Accra, Ghana;17(1):27–40.

[28] Okour A, Alkhateeb M, Amarin Z. Awareness of Danger Signs and Symptoms of Pregnancy Complication among Women in Jordan;118(1):11–4.

[29] Doctor HV, Findley SE, Cometto G, Afenyadu GY. Awareness of Critical Danger Signs of Pregnancy and Delivery, Preparations for Delivery, and Utilization of Skilled Birth Attendants in Nigeria;24(1):152–70. Available from: https://muse.jhu.edu/pub/1/article/496940.

[30] RCOG. Reduced Fetal Movements Green-Top Guideline No. 57;. Available from: https://www.rcog.org.uk/media/2gxndsd3/gtg_57.pdf.

[31] Kalisse T, Farrell A, Verling A, Rutherford E, Ravinder M, Khalid A, et al. National Clinical Practice Guideline: Reduced Fetal Movements.

[32] SASOG. REDUCED FETAL MOVEMENTS (VERSION 2.0 Final);. Available from: https://sasog.co.za/wp-content/uploads/2023/04/REDUCED-FETAL-MOVEMENTS_-2.0-final.pdf.

[33] Flenady V, Gardener G, Ellwood D, Coory M, Weller M, Warrilow K, et al. My Baby’s Movements: A Stepped-Wedge Cluster-Randomised Controlled Trial of a Fetal Movement Awareness Intervention to Reduce Stillbirths;129(1):29–41.

[34] Housseine N, Browne J, Maaløe N, Dmello BS, Ali S, Abeid M, et al. Fetal Movement Trials: Where Is the Evidence in Settings with a High Burden of Stillbirths?;130(3):241–3.

[35] Ghosh AK, Catelli DS, Wilson S, Nowlan NC, Vaidyanathan R. Multi-Modal Detection of Fetal Movements Using a Wearable Monitor; 103:102124.

[36] Ryo E, Kamata H, Yatsuki K. Correlation between Perinatal Abnormalities and Decreased Fetal Movement as Counted by a Fetal Movement Acceleration Measurement Recorder: A Prospective Cohort Study;169(2):656–62.

[37] Abrar A, Fairbrother N, Smith AP, Skoll A, Albert AYK. Anxiety among Women Experiencing Medically Complicated Pregnancy: A Systematic Review and Meta-Analysis;47(1):13–20.

[38] Gravensteen IK, Jacobsen EM, Sandset PM, Helgadottir LB, Ra destad I, Sandvik L, et al. Anxiety, Depression and Relationship Satisfaction in the Pregnancy Following Stillbirth and after the Birth of a Live-Born Baby: A Prospective Study;18(1):41.

[39] Kurki T, Hiilesmaa V, Raitasalo R, Mattila H, Ylikorkala O. Depression and Anxiety in Early Pregnancy and Risk for Preeclampsia;95(4):487. Available from: https://journals.lww.com/greenjournal/abstract/2000/04000/depression_and_anxiety_in_early_pregnancy_and_risk.3.aspx.

[40] Ding XX, Wu YL, Xu SJ, Zhu RP, Jia XM, Zhang SF, et al. Maternal Anxiety during Pregnancy and Adverse Birth Outcomes: A Systematic Review and Meta-Analysis of Prospective Cohort Studies;159:103–10.

[41] Dubuisson N, Diez Campa M, Ghosh AK, McAuliffe F, Nowlan N. Women’s perspectives on fetal movement monitoring in high and low stillbirth settings: a qualitative study. [Data set] Zenodo; 2025. 10.5281/zenodo.17572671

