## Supplementary Data for "Women’s perspectives on fetal movement monitoring in high and low stillbirth settings: a qualitative study"

### 1. Statistical analysis

Table 1: Mann-Whitney statistical tests indicate that reported ease of following FM monitoring advice was significantly higher in high stillbirth countries than in low stillbirth countries. No other factors had a significance effect on reported ease of following advice. 'Mean rank' represents the mean ranking of difficulty for each factor (a higher rank indicates increased difficulty), 'U' measures the difference between rank distributions, 'Z' is the standardised test value, and 'p-value' indicates the probability that the observed difference is due to chance.($*p \leq 0.05$,$** p \leq 0.01$)

| Factor | N | Mean Rank | U | Z | p-value |
|---|---|---|---|---|---|
| Stillbirth rate | | | | | |
| Low | 121 | 123.95 | 4482 | -3.37 | <0.001** |
| High | 100 | 95.33 | | | |
| Parity | | | | | |
| Primiparous | 95 | 119.98 | 5131 | -1.84 | 0.065 |
| Multiparous | 126 | 104.23 | | | |
| Complication | | | | | |
| Yes | 137 | 114.70 | 5247 | -1.12 | 0.264 |
| No | 84 | 104.97 | | | |
| Prior miscarriage | | | | | |
| Yes | 58 | 119.26 | 3842 | -1.72 | 0.085 |
| No loss | 156 | 103.13 | | | |
| Prior stillbirth | | | | | |
| Yes | 12 | 94.96 | 810 | -0.79 | 0.431 |
| No loss | 156 | 83.70 | | | |

Table 2: Kruskal Wallis statistical test indicate that there is a significant difference between rated ease of following advice between countries, however pairwise analysis is inconclusive. Age is shown to have a significant effect with participants aged 18-29 years old rating advice easier to follow than those 30-39 years old. ‘Mean rank’ represents the mean ranking of difficulty for each factor (a higher rank indicates increased difficulty), ‘H’ measures the difference between rank distributions, ‘df’ indicates the degrees of freedom, and ‘p-value’ indicates the probability that the observed difference is due to chance. ($*p \leq 0.05$,$**\ p \leq 0.01$)

| Factor | N | Mean Rank | H | df | p-value | Posthoc comparison |
|---|---|---|---|---|---|---|
| Country | | | | | | |
| Ireland | 46 | 122.99 | 15.45 | 6 | 0.017* | Posthoc tests non-significant |
| UK | 36 | 128.48 | | | | |
| USA | 39 | 120.91 | | | | |
| Kenya | 16 | 85.81 | | | | |
| South Africa | 48 | 89.14 | | | | |
| Zimbabwe | 24 | 117.06 | | | | |
| Other | 12 | 89.29 | | | | |
| Age (years) | | | | | | |
| 18–29 | 57 | 91.96 | 7.046 | 2 | 0.030* | 18–29<30–39 (p=0.027) |
| 30–39 | 144 | 117.63 | | | | |
| 40–49 | 20 | 117.55 | | | | |

Table 3: Mann-Whitney tests showed that participants from high SBR countries were significantly more likely to rate frequency of anxiety higher than participants in low SBR countries. Participants with pregnancy complications/risk factors, and those with experience of a prior stillbirth were also significantly more frequently anxious than those without. Parity and experience of a prior miscarriage were not shown to be statistically significant. 'Mean rank' represents the mean ranking of difficulty for each factor (a higher rank indicates increased difficulty), 'U' measures the difference between rank distributions, 'Z' is the standardised test value, and 'p-value' indicates the probability that the observed difference is due to chance. ($*p \leq 0.05$, $** p \leq 0.01$)

| **Factor** | **N** | **Mean Rank** | **U** | **Z** | **p-value** |
|---|---|---|---|---|---|
| Stillbirth rate | | | | | |
| Low | 122 | 109.11 | 5809 | -2.07 | 0.039* |
| High | 112 | 126.63 | | | |
| Parity | | | | | |
| Prima | 101 | 119.23 | 6541 | -0.36 | 0.722 |
| Multi | 133 | 116.18 | | | |
| Complication | | | | | |
| Yes | 146 | 126.72 | 5078 | -2.80 | 0.005** |
| No | 88 | 102.20 | | | |
| Prior miscarriage | | | | | |
| Yes | 63 | 126.40 | 4322 | -1.93 | 0.054 |
| No loss | 163 | 108.52 | | | |
| Prior stillbirth | | | | | |
| Yes | 14 | 120.64 | 698 | -2.51 | 0.012* |
| No loss | 163 | 86.28 | | | |

Table 4: Kruskal Wallis statistical test indicate that there are no statistically significant differences between rated frequency of anxiety between countries or age groupings. 'Mean rank' represents the mean ranking of difficulty for each factor (a higher rank indicates increased difficulty), 'H' measures the difference between rank distributions, 'df' indicates the degrees of freedom, and 'p-value' indicates the probability that the observed difference is due to chance. ($*p \leq 0.05$, $** \, p \leq 0.01$)

| **Factor** | **N** | **Mean Rank** | **H** | **df** | **p-value** | **Posthoc comparison** |
|---|---|---|---|---|---|---|
| Country | | | | | | |
| Ireland | 47 | 113.31 | 8.565 | 6 | 0.2 | Not applicable |
| UK | 36 | 109.75 | | | | |
| USA | 39 | 103.47 | | | | |
| Kenya | 18 | 115.39 | | | | |
| South Africa | 55 | 137.06 | | | | |
| Zimbabwe | 26 | 124.42 | | | | |
| Other | 13 | 102.50 | | | | |
| Age (years) | | | | | | |
| 18–29 | 64 | 129.46 | 3.022 | 2 | 0.221 | Not applicable |
| 30–39 | 150 | 112.74 | | | | |
| 40–49 | 20 | 114.93 | | | | |

### 2. Survey Questions

**About you**

1. * I:

   - am in my third trimester
   - was pregnant in the past year
   - Neither of the above apply

2. In what country do you live?

3. What is your age?

   - 17 or younger
   - 18-20 • 21-29 • 30-39
   - 40-49
   - 50 or older

4. How many times have you been pregnant?

   - 1 • 2 • 3
   - 4
   - 5+

5. How many babies have you given birth to?

   - 0 • 1 • 2 • 3
   - 4
   - 5+

6. Are/were any complications or risk factors associated with your current/recent pregnancy? Please tick all that apply.

   - Fetal growth restriction (small baby)
   - Pre-eclampsia and eclampsia
   - Gestational diabetes
   - High blood pressure
   - High BMI
   - Twins/triplets
   - Preterm birth
   - Other (please specify)

7. Did you ever have a miscarriage?

   - Yes
   - No

8. Did you ever have a stillbirth?

- Yes
- No

---

**Your baby's movements**

1. What advice have you received in your current pregnancy about monitoring your baby's movements?
2. How easy or hard do you find it to follow the advice you have received on monitoring your baby's movements?
    - Very easy
    - Easy
    - Somewhat easy
    - Neither easy nor difficult
    - Somewhat difficult
    - Difficult
    - Very difficult
    - N/a
3. Could you please tell us why?
4. How often do you feel worried or anxious about your baby's movements?
    - Always
    - Usually
    - Sometimes
    - Rarely
    - Never

### 3. Semi-structured Interview Questions

1. Which country do you live in?
2. How far along are you in your current pregnancy or when did you give birth?
3. How many times have you been pregnant?
4. What advice have you heard about monitoring fetal movements? How easy was it to follow this advice?
5. What are your thoughts and feelings about keeping track of baby's movements? During your pregnancy do/did you feel anxious about monitoring fetal movements?

### 4. Reflexivity Statement

The lead authors and majority of the co-authors are living and working in a high-income setting with low stillbirth rates. This may influence the lens through which the study was designed and the data analysed.
FM is a Professor of Obstetrics and Gynaecology in Ireland and therefore is familiar with fetal movement monitoring advice given in Ireland. The authors are currently or have previously worked on the development of a wearable fetal movement monitor and therefore have more knowledge about fetal movements than the average person.
Participants were not known previously to any of the authors.
